# Differential Results of Polygenic Risk Scoring for Multiple Sclerosis in European and African American Populations

**DOI:** 10.1101/2024.06.11.24308714

**Authors:** Cyprien A. Rivier, Seyedmehdi Payabvash, Hongyu Zhao, David A. Hafler, Guido J. Falcone, Erin E. Longbrake

**Affiliations:** Department of Neurology, Yale School of Medicine, New Haven, CT, USA; Yale Center for Brain and Mind Health, Yale School of Medicine, New Haven, CT, USA

## Abstract

**Importance:** The risk of multiple sclerosis (MS) is significantly influenced by polygenic inheritance. Polygenic risk scores (PRS) for MS can help identify high-risk individuals and stratify populations for clinical trials. However, most genome-wide association studies (GWAS) have been conducted in populations of European ancestry, raising questions about the accuracy of these PRS in other ancestries.

**Objective:** To determine whether a PRS for MS can effectively stratify individuals of non-European ancestries.

**Design, setting and participants:** This cross-sectional study utilized prospectively collected data from the All of Us Research Program (2018-2023). It included participants who had both whole genome sequencing and electronic health record (EHR) data.

**Exposure(s):** A PRS comprising 282 independent single nucleotide variants for MS, divided into quintiles.

**Main Outcome(s) and Measure(s):** Prevalence of multiple sclerosis ascertained through ICD-10 or SNOMED codes.

**Results:** In this study population, MS cases comprised 1.0% (327 cases) of the European population, 0.56% (183 cases) of the African population, and 0.46% (150 cases) of the Latino/admixed American population. In analyses adjusting for age, sex, and genetic principal components, the PRS associates with MS risk in the European population, with a 141% increase in the risk of MS for individuals in the highest compared to the lowest PRS quintile (OR: 2.41 [1.69-3.50], test-for-trend p<0.001). Similarly, the PRS appropriately partitions the Latino/admixed population into increasing MS risk groups (OR: 2.56 [1.45-4.78], test-for-trend p-value <0.001). However, it did not significantly stratify the African population into distinct MS risk categories (OR: 1.45 [0.95-2.25], test-for-trend p=0.10).

**Conclusions and Relevance:** A PRS for MS effectively stratified individuals of European and Latino/admixed ancestries but not African ancestry. This highlights the need for ancestry-specific PRS development to ensure accurate risk prediction across diverse populations, emphasizing the importance of including non-European groups in MS genetic research.

## Introduction

Multiple sclerosis (MS) is a chronic autoimmune disorder characterized by inflammation and demyelination in the central nervous system, leading to a range of neurological symptoms and disabilities^1^. The etiology of MS is complex, involving both genetic and environmental factors^1^. Advances in genomic technologies have significantly contributed to our understanding of the genetic architecture of MS, revealing that polygenic inheritance plays a crucial role in the disease’s development^2^. At present, 333 single nucleotide polymorphisms (SNPs) have been associated with risk of MS^3^, most are common variants that are associated with only modest increases in risk. However, the cumulative genetic burden of these loci may represent an important biomarker that is able to differentiate between those with high and low risk of future disease^4^.

Polygenic risk scores (PRS) have emerged as a powerful tool to quantify an individual’s cumulative genetic predisposition to complex diseases like MS^5^. PRS aggregate the effects of numerous genetic variants into a single score that can be used to predict disease risk. This method has shown promise in identifying high-risk individuals. Importantly, clinical trials evaluating lipid-lowering therapies have shown significant heterogeneity of treatment effects across categories of polygenic risk, with study participants at highest polygenic risk deriving larger benefits^6^. However, most genome-wide association studies (GWAS) that inform PRS development have predominantly involved populations of European ancestry^7^. This raises concerns about the applicability and accuracy of PRS in individuals of non-European ancestries, potentially leading to disparities in disease prediction and prevention efforts.

To address this gap, our study aims to evaluate the effectiveness of an MS-related PRS based on the known genetic architecture of the disease within different ancestry groups. The All of Us Research Program^8^ aims to enroll an ethnically diverse group of 1 million Americans to accelerate biomedical research and reduce health disparities. All of Us is generating whole genome sequencing data and incorporates complete electronic health records (EHR) data. Using All of Us, we sought to determine whether a PRS developed primarily from European ancestry data could effectively stratify individuals of African and Latino/admixed American (L/A) ancestries into distinct MS risk categories. Given the underrepresentation of non-European populations in genetic research, this study provides an essential assessment of PRS utility in diverse groups, thereby contributing to more equitable and accurate genetic risk assessment in MS.

## Methods

### Study design and inclusion criteria

We conducted a cross-sectional genetic study within the All of Us Research Program^8^, an ongoing study in the US that aims to enroll 1 million Americans aged 18 and older. All of Us does not focus on any particular disease or health status and emphasizes recruitment of groups that have been historically underrepresented in biomedical research. All of Us collects baseline health surveys at enrollment and uses several means to collect longitudinal health data, including continuous abstraction of EHR data in the form of billing codes, laboratory and medication data, radiology reports, and linkage with other data sources, such as national death indexes, pharmacy data, health care claims data, and geospatially linked environmental data. The All of Us data used for this analysis were collected between May 2018 and July 2023. Data from All of Us (https://www.researchallofus.org) is available to researchers by application. The All of Us institutional review board approved the study protocol for this study. All participants or their legally designated surrogates provided written informed consent. In the present nested study, we included participants with both whole genome sequencing (WGS) data and EHR data.

### Genomic data

DNA was isolated from blood and saliva samples obtained at dedicated research centers across the US, and WGS data were generated as described previously^9^. WGS data were quality controlled centrally using standardized pipelines that follow the most up-to-date standards in the field.

### Genetic ancestry ascertainment

Ancestry was ascertained centrally by the All of Us team using principal components analysis on WGS data by comparing participants with reference datasets from diverse populations. Each participant was assigned to: African, East Asian, South Asian, West Asian, European, L/A or Other. For the rest of the analysis, we only included participants belonging to the three largest ancestry groups: European, African, and L/A.

### Exposure ascertainment

Our exposure of interest was polygenic susceptibility to MS modeled through a PRS, a well-established tool in statistical genetics that estimates an individual’s genetic burden across numerous genetic risk variants.^10^ For a given study participant, the polygenic risk score is the sum of the product of the risk allele counts for each variant multiplied by the allele’s reported effect on MS. The score used genetic information on 282 independent genetic risk variants known to be associated with higher risk of MS. These genetic risk variants are single nucleotide polymorphisms with a minor allele frequency >1%, independent (r2, a measure of the correlation between variants, <0.1), biallelic (involve two alleles only) and associated with the risk of MS at genome-wide levels (P<5x10-8), following best practices for PRS generation.^3^

### Outcome assessment

Corresponding ICD-10, and Systemized Nomenclature of Medicine (SNOMED) codes were used to ascertain MS. Relative to baseline assessment, both prevalent and incident MS events were counted.

### Statistical analysis

We present discrete variables as counts (percentage [%]) and continuous variables as mean (standard deviation [SD]) or median (interquartile range [IQR]), as appropriate. Unadjusted comparisons were made using chi-square tests for discrete variables and t or ANOVA tests, as appropriate, for continuous variables. The three genetic ancestries considered (European, African, L/A) were randomly sampled to obtain three populations of equivalent sizes. In each population, we normalized the PRS (by subtracting the mean and dividing by the standard deviation) and divided it into quintiles to establish five risk categories (very low, low, intermediate, high and very high polygenic risk). To assess the relationship between the PRS and the risk of MS in each population, we used multivariable logistic regression models adjusting for age, sex, and the first four genetic principal components.

## Results

### Cohort characteristics

Of 413,457 participants included in the All of Us Research Program, 103,125 did not have EHR data, 137,179 did not have WGS data, and 173,153 were included in our analysis (mean age: 51.6 (SD: 17), 60% female). The distribution of genetic ancestry was as follows: 95,971 (55.4%) participants of European, 37,674 (21.8%) of African, 32,428 (18.7%) of L/A, 4,040 (2.3%) of East Asian, 2,365 (1.4%) of South Asian, and 675 (0.4%) of Middle Eastern genetic ancestry (Table 1). To obtain three populations of equal sizes, we randomly sampled 32,428 participants from the African and European populations to match the number of L/A participants.

**Table 1.**
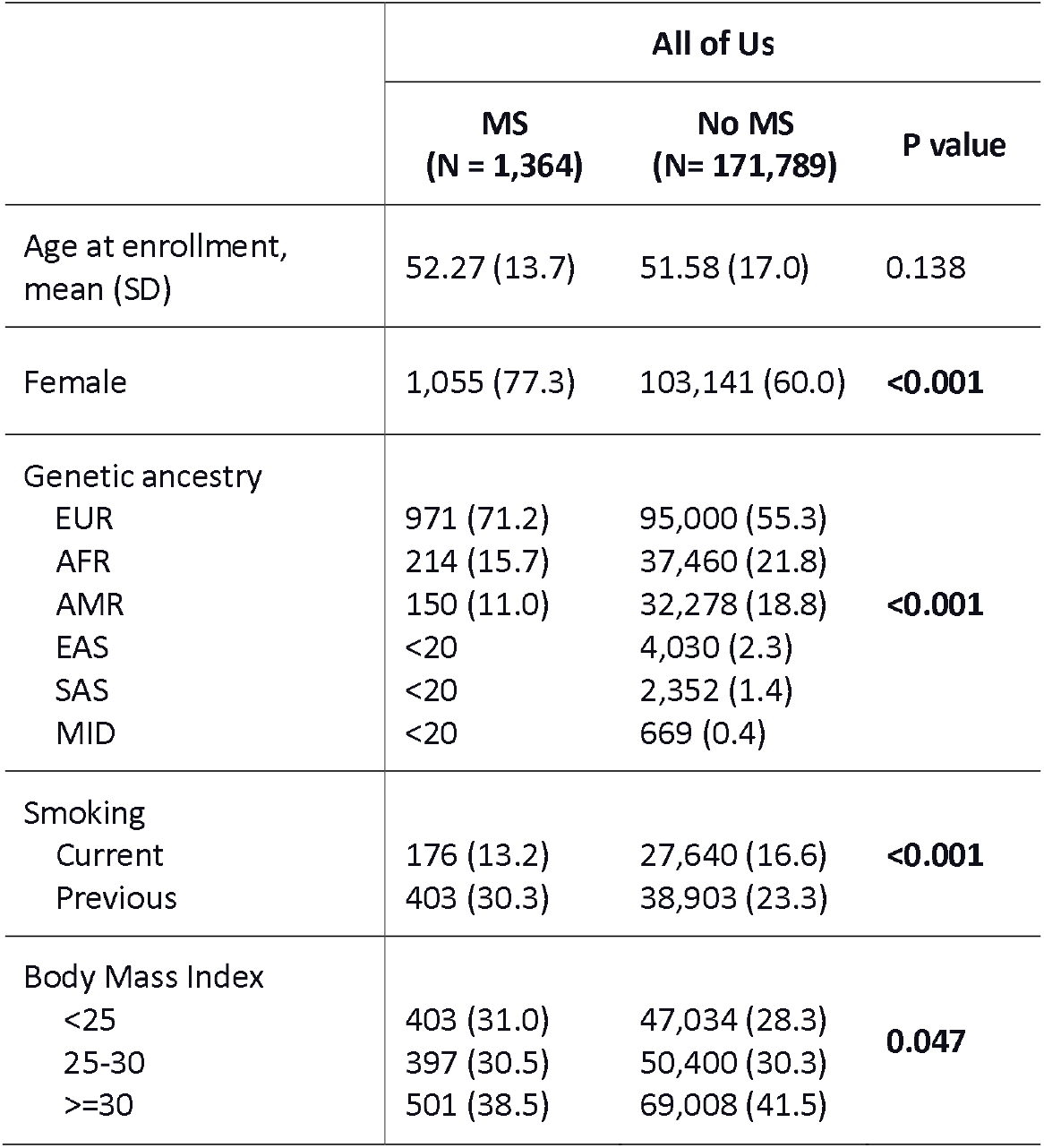
Cohort characteristics.

|  | All of Us |  |  |
| --- | --- | --- | --- |
|  | MS<br>(N = 1,364) | No MS<br>(N= 171,789) | P value |
| Age at enrollment,<br>mean (SD) | 52.27 (13.7) | 51.58 (17.0) | 0.138 |
| Female | 1,055 (77.3) | 103,141 (60.0) | **<0.001** |
| Genetic ancestry |  |  |  |
| EUR | 971 (71.2) | 95,000 (55.3) | **<0.001** |
| AFR | 214 (15.7) | 37,460 (21.8) |  |
| AMR | 150 (11.0) | 32,278 (18.8) |  |
| EAS | <20 | 4,030 (2.3) |  |
| SAS | <20 | 2,352 (1.4) |  |
| MID | <20 | 669 (0.4) |  |
| Smoking |  |  |  |
| Current | 176 (13.2) | 27,640 (16.6) | **<0.001** |
| Previous | 403 (30.3) | 38,903 (23.3) |  |
| Body Mass Index |  |  |  |
| <25 | 403 (31.0) | 47,034 (28.3) | **0.047** |
| 25-30 | 397 (30.5) | 50,400 (30.3) |  |
| >=30 | 501 (38.5) | 69,008 (41.5) |  |

### Unadjusted analysis

In each sample of 32,428 participants, there were 327 (1.0%) MS cases in the European group, 183 (0.56%) in the African group, and 150 (0.46%) in the L/A group. In the European population, the proportion of MS cases ranged from 0.66% in the lowest PRS quintile to 1.59% in the highest PRS quintile (p-value < 0.001 - Figure 1). In the African population, the proportion of MS cases did not increase consistently across PRS categories with the same strength. The smallest proportion (0.45%) occurred in the intermediate (3rd) PRS quintile, while the highest (0.82%) was in the highest PRS quintile (p-value: 0.06). In the L/A population, the proportion of MS cases increased by PRS quintile, ranging from 0.23% in the lowest quintile to 0.63% in the highest (p-value < 0.001).

**Figure 1.**
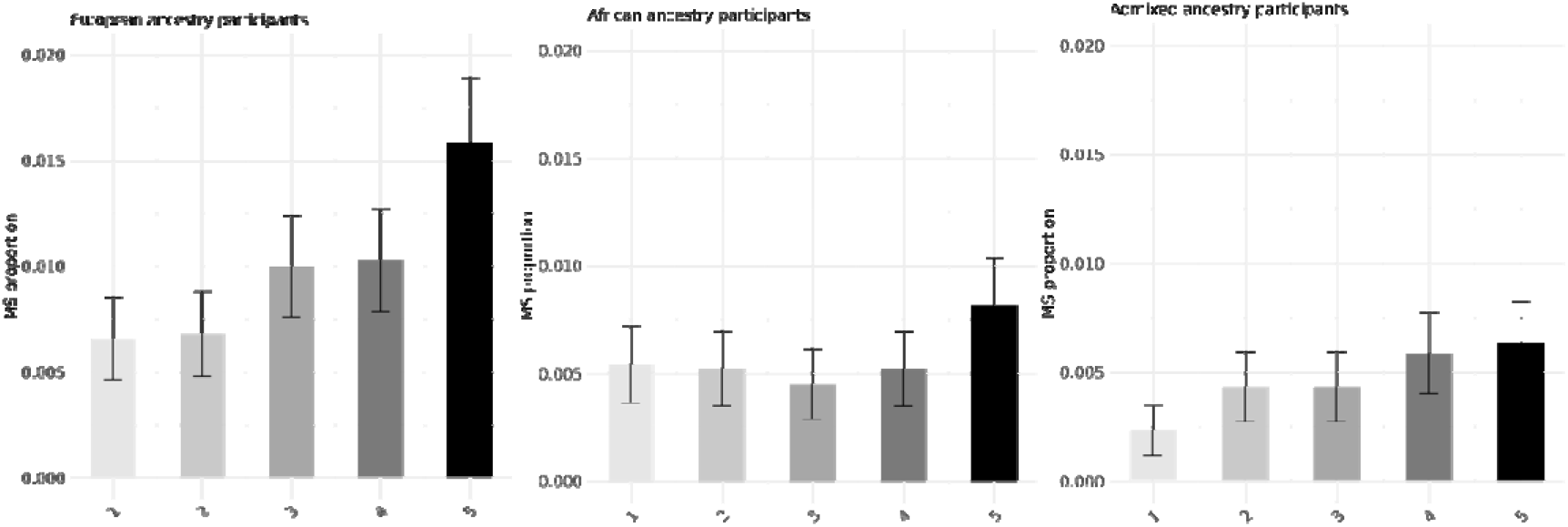
Proportions of MS cases by PRS quintile in European, African, and L/A ancestry populations.

### Adjusted analysis

In multivariable logistic regression adjusting for age, sex, and genetic principal components, we found that the PRS provided effective risk stratification in the European population. The odds of MS increased by 51% (Odds Ratio: 1.51, 95% CI: [1.03-2.25]) in the intermediate PRS quintile and by 141% (OR: 2.41 [1.69-3.50]) in the highest PRS quintile compared to the lowest (test for trend p-value: <0.0001 - Figure 2). In the African population, the PRS did not significantly stratify participants into different MS risk categories (test-for-trend p-value: 0.10). Conversely, in the L/A population, the PRS effectively stratified risk across several categories. The odds of MS increased by 85% (Odds Ratio: 1.85, 95% CI: [1.00-3.56]) in the intermediate PRS quintile and by 156% (OR: 2.56 [1.45-4.78]) in the highest quintile compared to the lowest (test-for-trend p-value: 0.0004).

**Figure 2.**
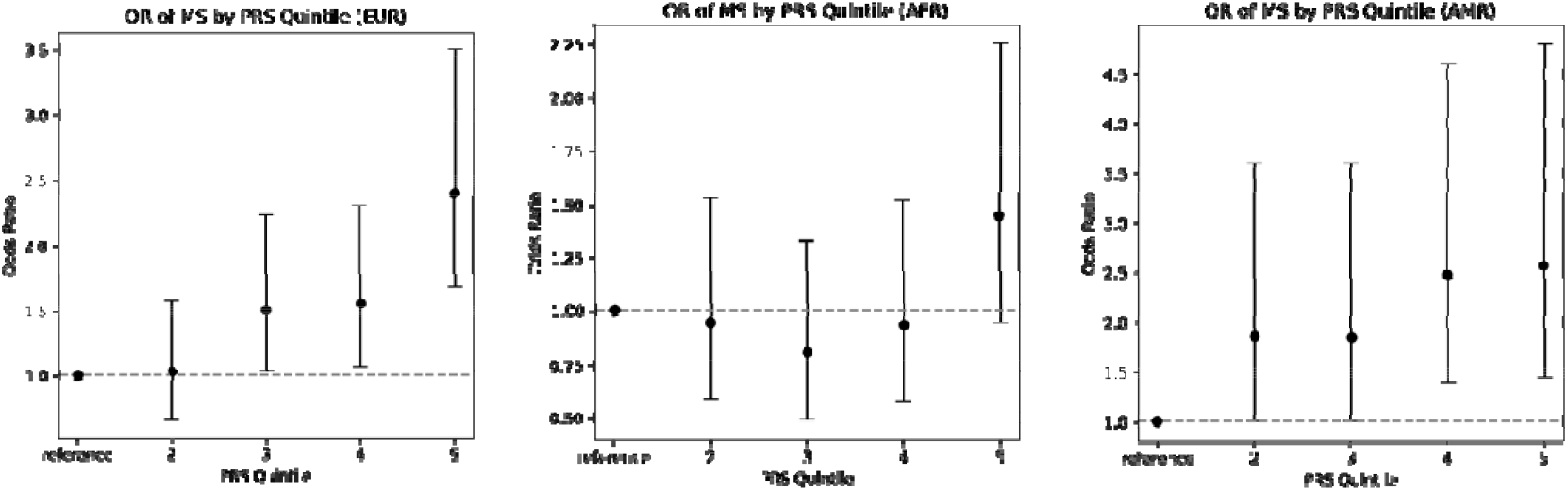
Adjusted odds ratios of MS diagnosis by PRS quintile in European, African, and L/A ancestry populations.

## Discussion

In this study, we evaluated the effectiveness of a PRS for MS across different genetic ancestries using data from the All of Us Research Program. Our findings reveal significant disparities in the utility of the PRS among European, African, and L/A populations, highlighting the necessity for tailored genetic risk prediction tools in diverse populations.

The PRS effectively stratified MS risk in European and L/A individuals, with higher PRS quintiles corresponding to significantly increased odds of MS. This confirms the robustness of PRS derived from predominantly European GWAS data for these groups. Specifically, individuals in the highest PRS quintile had significantly elevated odds of developing MS compared to those in the lowest quintile, consistent with previous studies demonstrating the potential of PRS in risk stratification and early disease identification^2,11^.

Conversely, the PRS did not significantly stratify MS risk among individuals of African ancestry. This discrepancy may stem from several factors. Primarily, the limited representation of African populations in existing GWAS likely results in PRS that are less applicable to this group^12^. The genetic variants contributing to MS risk in Europeans may not have the same effect in African populations, indicating differences in the genetic architecture of MS across ancestries^13^. Additionally, gene-environment interactions could play a role; genetic variants might interact with environmental factors differently across populations, affecting disease risk in ways that a PRS developed in one population cannot capture^14^. For instance, lifestyle, diet, and exposure to certain environmental factors can vary widely between populations, influencing how genetic predispositions manifest as disease. Moreover, African populations possess greater genetic diversity compared to European populations^15,16^. This diversity means the distribution of genetic variants is broader, including many that may be rare or absent in European populations. As a result, a PRS developed using European data may not account for the full range of genetic factors influencing MS risk in African populations. This genetic diversity complicates the direct application of PRS across different ancestries, necessitating the development of population-specific PRS to ensure accurate and equitable risk prediction.

Our study also highlights the importance of utilizing large, diverse datasets like the All of Us Research Program to evaluate genetic tools’ performance across populations. Such datasets are invaluable for understanding complex diseases’ genetic basis in diverse groups and developing inclusive genetic risk prediction models.

This study has several limitations that warrant consideration. Firstly, the absolute risk of MS in the population is low, which may limit the practical utility of the PRS as a stand-alone screening tool. Given the multifactorial nature of MS, a comprehensive screening approach that incorporates genetic, environmental, and lifestyle factors may be necessary to more accurately identify individuals at risk. Additionally, the performance of the PRS in the All of Us dataset was lower compared to its performance in other large datasets, such as the UK Biobank, even when considering individuals of European ancestry only^2^. This discrepancy suggests that specific factors or characteristics unique to the All of Us dataset, beyond genetic ancestry, may influence PRS performance. Factors such as differences in data collection methods, population heterogeneity, or unmeasured environmental influences could contribute to this variation.

In conclusion, while the PRS for MS is effective in European and L/A populations, it is less effective for African ancestry individuals. This emphasizes the urgent need for inclusive genetic research and the development of ancestry-specific PRS to ensure equitable benefits from genetic risk prediction and personalized healthcare. Future studies should focus on expanding genetic diversity in research cohorts, improving underrepresented populations’ representation in GWAS, and refining PRS methodologies to enhance their applicability across all populations, ultimately advancing the goal of personalized medicine for all.

## Data Availability

Data from All of Us (https://www.researchallofus.org) is available to researchers by application.

## Data availability statement

This study used data from the All of Us Research Program’s Controlled Tier Dataset C2022Q4R9 and Registered Tier Dataset R2022Q4R9, available to authorized users on the Researcher Workbench via www.allofus.nih.gov. All data access and analyses were conducted within a secure informatic workspace provided by the National Institutes of Health.

